# Prevalence and factors associated with reporting of sexual violence among secondary school adolescents in Ibadan Metropolis, Nigeria: A cross-sectional study

**DOI:** 10.64898/2026.04.08.26344946

**Authors:** Halimat O. Olaniyan, Adesola O. Olumide

## Abstract

**Background:** Sexual violence (SV) is a major public health problem with far-reaching consequences; however, adolescent survivors rarely seek help. This underestimates the prevalence of SV and undermines prevention and response efforts. This study was conducted to determine the prevalence and reporting of sexual violence among adolescents in Ibadan, Nigeria.

**Methods:** Between September and October 2021, a cross-sectional study was conducted among 360 in-school adolescents in Ibadan South-West local government area, Nigeria. Adolescents were selected using multi-stage sampling. Information on experience and reporting of sexual violence was obtained with the aid of an interviewer-administered questionnaire. Data were analysed using descriptive and inferential statistics at p≤0.05 level.

**Results:** Respondents’ mean age was 14.6±1.7 years, and 50% were female. Thirty-five per cent reported at least one incident within the past 12 months. Forms of sexual violence experienced included unwanted sexual touch (25.6%), forced sex (19.2%), attempted rape (15.2%), and suggestive comments (9.6%). Seventy per cent of adolescents who experienced sexual violence did not report to anyone; reasons included fear of getting in trouble (46.6%), thinking it was not a problem (31.8%), feeling it was their fault (30.7%), and embarrassment for self/family (27.3%). Adolescents who were closer to their mothers and younger adolescents were more likely to report their experience of sexual violence (p=0.006 and p=0.038, respectively).

**Conclusion:** Sexual violence is common among in-school adolescents in Ibadan, yet reporting remains low. This study highlights the need to strengthen prevention and address barriers to reporting among adolescent survivors.

**Key Message:** *What is already known on this topic:* Sexual violence has physical, psychological, and social consequences on the health and well-being of adolescent survivors, and low levels of reporting and help-seeking contribute to these consequences.

*What this study adds:* This study provides evidence on the prevalence and reporting patterns of sexual violence among adolescents in Ibadan, Nigeria. It highlights key barriers and facilitators of reporting.

*How this study might affect research, practice, or policy:* This study provides information about factors at individual, relationship, societal and policy levels that are associated with reporting and help-seeking among adolescent survivors of sexual violence in Ibadan, Nigeria. This highlights the importance of training stakeholders, such as parents, teachers, health workers, other caregivers and the adolescents themselves, on responding to sexual violence experience and reporting. It underscores the need for improved adolescent-friendly services, policy implementation and collaboration across families, schools, communities and states to address sexual violence.

## Background

Sexual violence is a major public health problem, with physical, social and psychological consequences (1–4). The World Health Organization (WHO) describes sexual violence as, “any sexual act, attempt to obtain a sexual act, unwanted sexual comments or advances, or acts to traffic, or otherwise directed, against a person’s sexuality using coercion, by any person, regardless of their relationship to the victim, in any setting, including but not limited to home and work” (2). Sexual violence includes both contact and non-contact forms, such as harassment, exploitation, online sexual abuse, completed and attempted forced penetration (rape), as well as other unwanted sexual acts involving force or threat of force, or situations where the victims cannot give consent due to age, disability, authority dynamics or under the influence of alcohol or other drugs (2,5–8).

Adolescents are particularly vulnerable to sexual violence due to limited knowledge and skills (9), stage of development (10), power imbalances and societal norms (11–13). Available global data reveal that the prevalence of adolescent SV varies widely within and between countries. Under-reporting, differences in methodology used, and paucity of nationally representative current data contribute to the differences in rates of adolescent sexual violence. A report by UNICEF (2025) documented that an estimated 650 million women and girls (approximately one in five) have experienced sexual violence during childhood, while between 410 and 530 million men and boys (around one in seven) report having experienced sexual violence before the age of 18 (14). In Nigeria, the Violence Against Children Survey (VACS) found that approximately 25% of girls and 10–11% of boys experienced sexual violence before the age of 18 (15).

Sexual violence against adolescents remains a global yet underreported problem, especially in Sub-Saharan Africa, where many victims are reluctant to disclose their experiences or seek help (4,16,17). Adolescents often experience sexual violence in familiar places, such as homes, workplaces, schools, religious centres, health facilities, or markets and perpetrators are often known to the survivors(2,18,19). Common perpetrators include friends, romantic partners, neighbours, family members and relatives (15,20). In schools, increased interaction with peers and limited supervision by adults may increase the risk of peer-on-peer victimisation (21). This can affect their education and extend into adulthood to affect their overall well-being (1).

Various factors are associated with experience and reporting of sexual violence at the individual, relationship, community and institutional levels. Non-reporting is often rooted in social and cultural norms that promote violence (19,22,23). Many survivors do not report their experiences to authorities or make informal disclosures to anyone due to denial, fear, shame, guilt, or a lack of trust in formal response systems (4,5,24–26). Adolescents may not classify acts of sexual violence correctly, thereby minimising or underreporting their experiences (27). Survivors are often female; therefore, interventions have focused mainly on them. However, studies have shown that males also experience violence, and there are indications that males are less likely to report it than females (27,28).

Most research in Nigeria and other parts of Sub-Saharan Africa focuses on the experiences of older individuals, with less emphasis on adolescent populations, and there has been limited research on adolescent sexual violence and barriers to disclosure (13,29,30). Existing measures, such as the provision of sexual assault referral centres, laws prohibiting violence, and awareness by various non-governmental organisations, have been put in place to address violence. However, the implementation of these measures is insufficient, and underreporting persists for various reasons.

This study aims to provide information to address the problem of under-reporting by assessing the prevalence and factors associated with reporting sexual violence among adolescents aged 10–19 in Ibadan, Nigeria, specifically; i) to determine the prevalence of sexual violence among adolescents, ii) to determine self-reported consequences of sexual violence on survivors iii) to describe reporting practices among survivors of sexual violence iv) to determine factors associated with reporting of sexual violence by adolescent survivors. “Reporting” in this study includes formal reporting (e.g., to police/health workers) or any disclosure (to friends/family)

**Figure 1:**
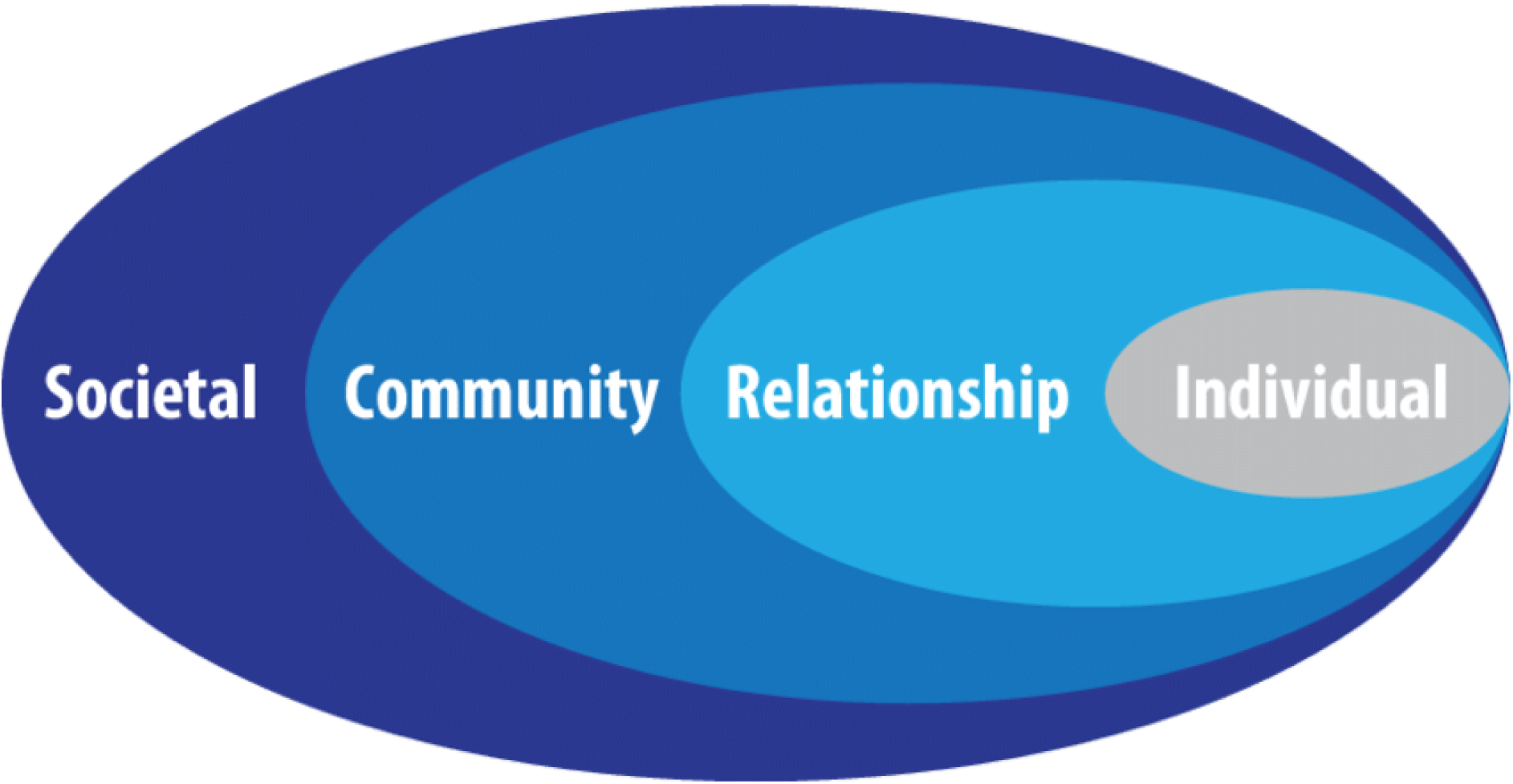
The socio-ecological model (2)

### Theoretical framework

The social-ecological model was used in this study to inform the selection of study variables and interpretation of findings on how factors at multiple levels influence sexual violence experiences and reporting among adolescents. The model is commonly used to understand how factors at individual, relationship, community and societal levels serve as risk and protective factors for violence experience and response (13,31–34).

Individual-level factors such as demographic characteristics and biological factors influence how individuals behave and increase their tendencies to become victims or perpetrators of sexual violence (13). At the **individual level**, sociodemographic characteristics such as age, gender, class and religion were assessed to measure the likelihood of sexual violence experience and reporting among adolescents

At the **relationship level,** sexual violence occurs within the various contexts in which adolescents interact with others. In this study, relationship-level factors considered include living arrangements, parental marital status, and reported closeness to parents or guardians, which may influence experience and reporting.

At the **Community levels,** we explored settings such as schools and neighbourhoods. Considering the amount of time adolescents spend in school environments, markets and neighbourhoods, these settings may serve as sites of both vulnerability and protection.

At the **societal level,** factors such as societal norms and culture, economic factors, laws, policies and educational factors affect sexual violence as identified in this study, providing a context in which other factors operate.

## Materials and Methods

### Study Design and Setting

A cross-sectional study was conducted among in-school adolescents aged 10 to 19 in Ibadan South-West Local Government Area (LGA), Oyo State, Nigeria. Ibadan South-West is one of the five urban LGAs in the Ibadan metropolis and has about 36 government-owned secondary schools, according to records from the Oyo State Ministry of Education

### Study Population and Eligibility Criteria

The study included male and female adolescents aged 10-19 attending selected government-owned secondary schools in Ibadan, South-west Nigeria, who were present during data collection and provided assent with parental consent (for those aged 10–17 years) or informed consent (for those aged 18–19 years). Participants were selected from junior secondary (JSS2 and JSS3) and senior secondary (SS2 and SS3) classes. Adolescents in JSS1 and SS1 were excluded because they had not yet resumed classes at the time of recruitment and data collection; the session had just begun, and students in these classes were new and had not yet been placed in schools, as they were just starting either the Junior School or Senior School.

### Sampling Technique

A multi-stage sampling approach was used. Ibadan South-West was randomly selected from the five urban LGAs within Ibadan. Ten of the thirty-six public secondary schools in Ibadan South-West were then selected using simple random sampling (balloting without replacement) In each selected school, one class arm was selected from each eligible level (Junior Secondary School [JSS] and Senior Secondary School [SSS]) using simple random sampling by balloting. Participants were selected from the designated class arms using simple random sampling. Within the selected class arm, a list of all students (class register) was obtained, and participants were selected using simple random sampling. Each student in the list was assigned a number, and numbers were randomly generated to select participants for the study.

The sample size was calculated using the Leslie Kish (35), formula for single proportions:

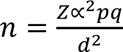

where *n* is the minimum sample size, *Z* is the standard normal deviate at 95% confidence level (1.96), *p* is the estimated prevalence of sexual violence the prevalence of sexual violence among male and female adolescents (34.9%) reported in a study conducted by Ajuwon et al. (2011) (22) and *d* is the margin of error (0.05).

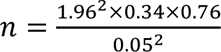

Substituting these values gave a minimum sample size of 350. To account for a 10% non-response rate, the sample size was adjusted to 385. A total of 360 adolescents completed the survey, giving a response rate of 93.5%.

### Study instrument

Data were collected using a semi-structured interviewer-administered questionnaire adapted from previous studies and relevant literature. The questionnaire assessed socio-demographic characteristics, experiences of sexual violence (lifetime and past 12 months), self-reported consequences, reporting and help-seeking behaviours. The instrument was reviewed for content validity through literature review and peer assessment. A pretest was done in a comparable school, and questions were refined for clarity. Internal consistency reliability testing yielded a Cronbach’s alpha coefficient of 0.943.

### Data Collection Procedure

Data was collected through interviewer-administered questionnaires to enhance data quality. Questionnaires were administered by trained interviewers, all postgraduate students of Child and Adolescent Health, who were familiar with working with adolescents. They were trained for two days before data collection commenced, with one day conducted virtually via Zoom and one day in person. Data collection was conducted over three weeks. The field supervisor reviewed all completed questionnaires, and regular meetings were held with the team to facilitate discussion, resolve issues, and allow the research team to express their views on the data collection.

The questionnaires were administered in a private location within the schools, such as the library for some schools and an open space (on the field) for others. Guidelines for obtaining information on violence safely, as recommended by the World Health Organisation (36), were followed; referral information was also provided for the participants with contact details and addresses of where to seek help within their communities. We also provided the contact of the lead investigator in case they needed help in connecting with help centres. As the study was conducted in 2021, when COVID-19 pandemic controls were still in force, COVID-19 control protocols, such as the use of facemasks and hand sanitisers, were duly observed during data collection.

### Patient and Public Involvement

The participants and other stakeholders were involved in the design of the study instrument through various reviews and consultations, as well as in the dissemination of the study findings.

### Data Analysis and Management

The data obtained were collated, cleaned, and analysed using IBM SPSS version 23. Data cleaning involved checking accuracy, handling missing values, and organising variables. Questionnaires were serially numbered for easy entry. Descriptive data were summarised using frequencies, percentages, and means for relevant variables. Bivariate analysis: the chi-square test was used to assess associations.

### Study variables

The primary outcome variable was **reporting of sexual violence**, defined as disclosure of an experience of sexual violence to any individual or authority, including family members, friends, teachers, health workers, or law enforcement agents. Selected independent variables aligned with the socioecological model and information from previous literature (31,32). Variables included socio-demographic characteristics such as age, current relationship status, class, employment status, closeness to parent/guardian, parent marital status, living arrangement, religion, and ethnicity. Outcome variables included experience covering lifetime and 12-month prevalence of sexual violence, self-reported consequences, reporting behaviours and professional help-seeking

### Ethical considerations

Ethical approval was obtained from the ethical review board of the Oyo State Ministry of Health. Permission was also obtained from the Ministry of Education and the school principals. Informed consent was obtained from adolescents older than 18, and assent was obtained from those under 18. In addition, written informed consent was obtained from parents/guardians of adolescents younger than 18 who participated in the study, as they were given forms stating that they were participating in a health study prior to data collection. The principles of voluntariness, beneficence, non-maleficence, and confidentiality were strictly followed. Participants were also provided information on available prevention and response services and linked to these services on request.

## Results

### Socio-demographic characteristics of respondents and their family characteristics

Of the 360 in-school adolescents interviewed, two-thirds (64.7%) were in the 14-16 age group, with a mean age of 14.6±1.7 years; (67.8% were Christians, 69.7% live with both parents, and 79.4% of parents are married. About half of the respondents reported being close or very close to their father (49.1%), while a higher proportion reported being close or very close to their mother (60.0%). Forty-three percent, were engaged in work for financial gain, 46.7% had ever had a romantic partner, while 26.4% currently had a romantic partner (Table 1).

**Table 1:** Socio-demographic characteristics of respondents and their family characteristics.

| **Characteristics** | **Frequency** | **Percentage** |
| --- | --- | --- |
| **Age (in years)** |  |  |
| 14-16 | 233 | 64.7 |
| 10-13 | 85 | 23.6 |
| 17-19 | 42 | 11.7 |
| **Gender** |  |  |
| Male | 180 | 50.0 |
| Female | 180 | 50.0 |
| **Religion** |  |  |
| Christianity | 244 | 67.8 |
| Islam | 113 | 31.4 |
| Traditional | 3 | 0.8 |
| **Class** |  |  |
| Senior secondary school | 180 | 50.0 |
| Junior secondary school | 180 | 50.0 |
| **Ethnic group** |  |  |
| Yoruba | 304 | 84.4 |
| Igbo | 45 | 12.5 |
| Hausa | 6 | 1.7 |
| Others | 5 | 1.4 |
| **Living arrangement** |  |  |
| Both parent | 251 | 69.7 |
| Mother | 51 | 14.2 |
| Guardian | 30 | 8.3 |
| Father | 28 | 7.8 |
| **Parents' Marital Status** |  |  |
| Married | 286 | 79.4 |
| Separated | 31 | 8.6 |
| Divorced | 19 | 5.3 |
| Never married | 10 | 2.8 |
| Widowed | 14 | 3.9 |
| **Engagement in work for financial gains** |  |  |
| No | 204 | 56.7 |
| Yes | 156 | 43.3 |
| **Relationship status** |  |  |
| Ever had a romantic partner | 168 | 46.7 |
| Currently have a romantic partner | 95 | 26.4 |
| **Closeness to father/male guardian** |  |  |
| Not close | 171 | 47.5 |
| Close | 138 | 38.3 |
| Very close | 39 | 10.8 |
| Never had a relationship with them | 12 | 3.3 |
| **Closeness to mother/female guardian** |  |  |
| Close | 150 | 41.7 |
| Not close | 131 | 36.4 |
| Very close | 66 | 18.3 |
| Never had a relationship with them | 13 | 3.6 |

### Prevalence of sexual violence among respondents

Forty two percent of the adolescents reported a lifetime experience of at least an act of sexual violence. Commonly reported forms of sexual violence ever experienced were unwanted sexual touching 66 (18.3%), coercion to look at a sexual act or pornographic materials 39 (10.8%), attempts to forcefully have sex and 38 (10.6%) and forced sex 35 (9.7%).

In the 12 months preceding the study, 125 (34.7%) of all 360 adolescents experienced at least one act of sexual violence. Common forms of sexual violence experienced in the preceding 12 months were, unwanted sexual touching 32 (25.6%), had experienced forced sex 24 (19.2%) attempts to have sex forcefully 19 (15.2%) and unwanted suggestive sexual comments 12 (9.6%). (Table 2).

**Table 2:**
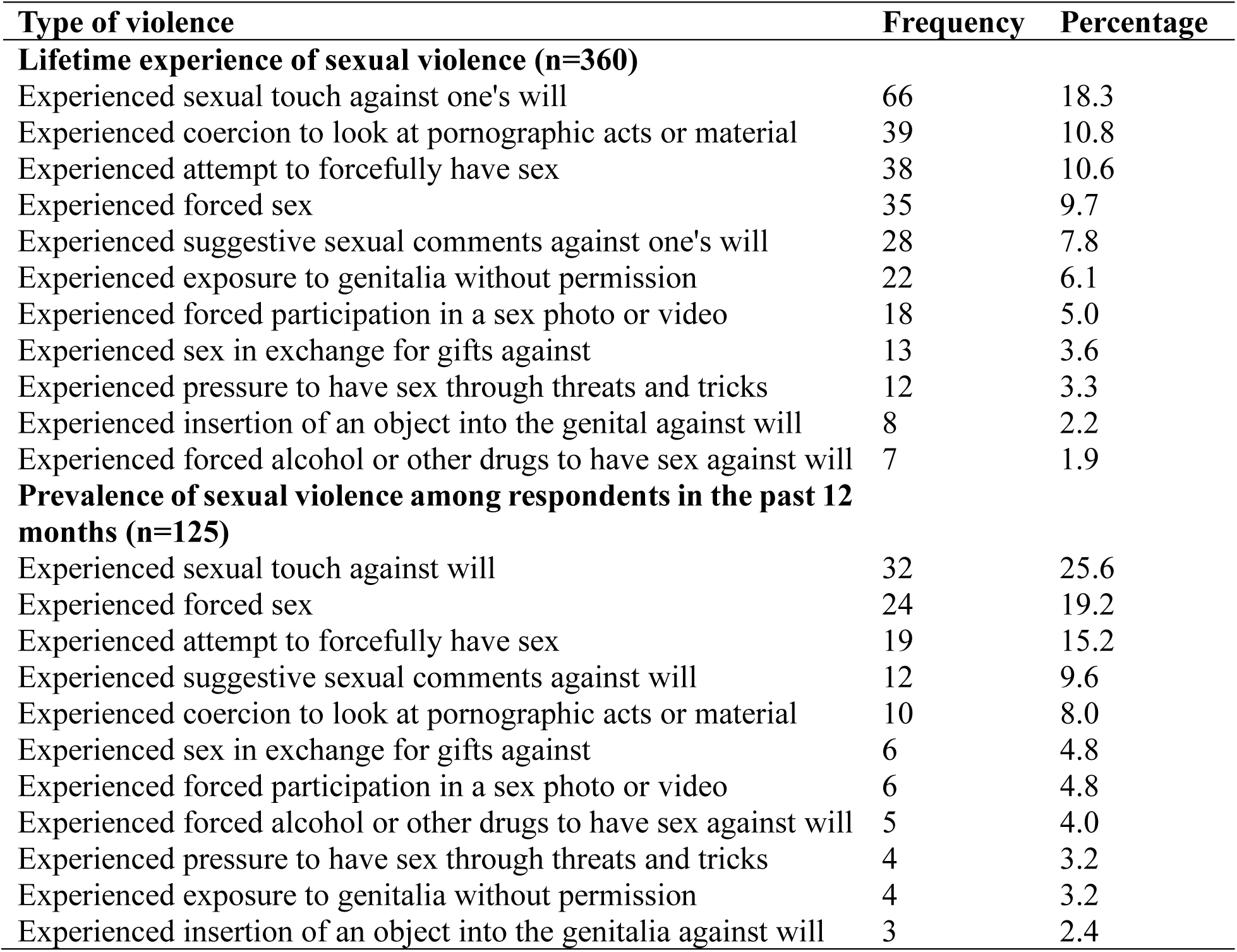
Lifetime and twelve-month prevalence of all acts of sexual violence among adolescents in Ibadan metropolis.

| Type of violence | Frequency | Percentage |
| --- | --- | --- |
| **Lifetime experience of sexual violence (n=360)** |  |  |
| Experienced sexual touch against one's will | 66 | 18.3 |
| Experienced coercion to look at pornographic acts or material | 39 | 10.8 |
| Experienced attempt to forcefully have sex | 38 | 10.6 |
| Experienced forced sex | 35 | 9.7 |
| Experienced suggestive sexual comments against one's will | 28 | 7.8 |
| Experienced exposure to genitalia without permission | 22 | 6.1 |
| Experienced forced participation in a sex photo or video | 18 | 5.0 |
| Experienced sex in exchange for gifts against | 13 | 3.6 |
| Experienced pressure to have sex through threats and tricks | 12 | 3.3 |
| Experienced insertion of an object into the genital against will | 8 | 2.2 |
| Experienced forced alcohol or other drugs to have sex against will | 7 | 1.9 |
| **Prevalence of sexual violence among respondents in the past 12 months (n=125)** |  |  |
| Experienced sexual touch against will | 32 | 25.6 |
| Experienced forced sex | 24 | 19.2 |
| Experienced attempt to forcefully have sex | 19 | 15.2 |
| Experienced suggestive sexual comments against will | 12 | 9.6 |
| Experienced coercion to look at pornographic acts or material | 10 | 8.0 |
| Experienced sex in exchange for gifts against | 6 | 4.8 |
| Experienced forced participation in a sex photo or video | 6 | 4.8 |
| Experienced forced alcohol or other drugs to have sex against will | 5 | 4.0 |
| Experienced pressure to have sex through threats and tricks | 4 | 3.2 |
| Experienced exposure to genitalia without permission | 4 | 3.2 |
| Experienced insertion of an object into the genitalia against will | 3 | 2.4 |

### Perpetrators of sexual violence against adolescent survivors in the Ibadan metropolis in the past 12 months

Among the 125 adolescents who had experienced sexual violence in the past 12 months, the most frequently reported perpetrator was a romantic partner 36.0%, friend 16.0%, neighbour 14.4%, classmate/school mate 12.8%. Less than five percent mentioned that the perpetrator was a stranger..

### Location where sexual violence occurred in the past 12 months

Information on the location where adolescents experienced the act of sexual violence is presented in Table 3. The survivors’ home was the most commonly reported location of sexual violence (46.4%), followed by perpetrators’ home (20.0%), shop or market (13.6%), and school (14.4%).

**Table 3:** Perpetrators, location, and self-reported consequences of sexual violence among survivors in Ibadan metropolis in the past 12 months.

| **Perpetrators, location, and self-reported consequences (n=125)** | **Frequency** | **Percent** |
| --- | --- | --- |
| **Perpetrators of sexual violence (n=125)** |  |  |
| Romantic partner | 45 | 36.0 |
| friend | 20 | 16.0 |
| neighbor | 18 | 14.4 |
| classmate/schoolmate | 16 | 12.8 |
| stranger | 6 | 4.8 |
| uncle | 5 | 4.0 |
| brother | 4 | 3.2 |
| teacher | 3 | 2.4 |
| father | 3 | 2.4 |
| employer | 2 | 1.6 |
| aunt | 1 | 0.8 |
| sister | 1 | 0.8 |
| stepfather | 1 | 0.8 |
| **The location where sexual violence occurred** |  |  |
| My home (Survivor's home) | 58 | 46.4 |
| Perpetrator's home | 25 | 20.0 |
| School | 18 | 14.4 |
| Shop/market | 17 | 13.6 |
| Someone else's home | 7 | 5.6 |
| **Self-reported consequences of sexual violence (n=125)\*** |  |  |
| Fear | 54 | 43.2 |
| Physical injury | 30 | 24.0 |
| Guilt | 26 | 20.8 |
| STI | 13 | 10.4 |
| Depression | 11 | 8.8 |
| Self-harm | 9 | 7.2 |
| Abortion | 1 | 0.8 |
| Unintended pregnancy | 1 | 0.8 |

\*Multiple responses

### Self-reported consequences of sexual violence among survivors in the past 12 months

Information on self-reported consequences experienced by adolescents following an incident of sexual violence in the past 12 months is presented in Table3. Common consequences experienced were fear (43.3%), physical injury 24%, and guilt (20.8%).

### Reporting of sexual violence in the past 12 months

Out of 125 respondents who experienced sexual violence in the past 12 months, the majority, 88 (70.4%), did not report their experience of sexual violence to anyone. At the **relationship level**, among the 37 respondents who reported to someone, most disclosed to their mothers (48.6%), followed by friends (21.6%). Most of those who disclosed their experience reported that the help received was helpful (86.5%). The help provided included advice 56.8%, warning the perpetrator (35.1%), and warning the respondent to be careful (8.1%). At the **individual level**, reasons for non-disclosure included fear of getting into trouble (46.6%), not perceiving the incident as a problem (31.8%), and feeling that it was their fault (30.7%) (Table 4).

**Table 4:** Reporting of sexual violence experienced by adolescent survivors in the 12 months before this study.

| **Reporting of sexual violence** | **Frequency** | **Percentage (%)** |
| --- | --- | --- |
| **Reported last experience of sexual violence (n=125)** |  |  |
| Yes | 37 | 29.6 |
| No | 88 | 70.4 |
| **The person reported to (n=37) \*** |  |  |
| Mother | 18 | 48.6 |
| Friend | 8 | 21.6 |
| Father | 5 | 13.5 |
| Teacher | 4 | 10.8 |
| Brother | 4 | 10.8 |
| Sister | 1 | 2.7 |
| Neighbor | 3 | 8.1 |
| Romantic partner | 2 | 5.4 |
| **Were they helpful (n=37)** |  |  |
| Yes | 32 | 86.5 |
| No | 5 | 13.5 |
| **Help provided (n=37)** |  |  |
| Advice | 21 | 56.8 |
| Warned the perpetrator | 13 | 35.1 |
| Warned me to be careful | 3 | 8.1 |
| **Reasons for not reporting (n=88)\*** |  |  |
| Afraid of getting in trouble | 41 | 46.6 |
| Did not think it was a problem | 28 | 31.8 |
| Felt it was my fault | 27 | 30.7 |
| Embarrassed for self/family | 24 | 27.3 |
| Perpetrator threatened me | 10 | 11.4 |
| Afraid of being abandoned | 1 | 1.1 |

\*Multiple responses

### Association between socio-demographic factors and reporting of sexual violence

The socio-demographic factors associated with reporting of sexual violence by adolescent survivors are reported in Table 5.1. At the **individual level** of the socio-ecological model, age and religion were significantly associated with reporting, while sex and class were not statistically significant. Reporting of sexual violence differed significantly by age group (χ² = 6.56, p = 0.038), with adolescents aged 10–13 years reporting more frequently (46.7%) compared with those aged 14–16 years (22.2%) and 17–19 years (35.7%). A greater proportion of Muslim adolescents (42.9%) reported their experience of sexual violence compared with Christians (22.0%), and this difference was statistically significant (χ² = 8.22, p = 0.016).

**Table 5.1:** Association between socio-demographic factors and reporting of sexual violence.

| Variables | Yes | No | Total | X<sup>2</sup> | P value |
| --- | --- | --- | --- | --- | --- |
| **Age** |  |  |  |  |  |
| 10-13 | 14 (46.7%) | 16 (53.3%) | 30 | 6.56 | 0.038 |
| 14-16 | 18 (22.2%) | 63 (77.8%) | 81 |  |  |
| 17-19 | 5(35.7%) | 9 (64.3%) | 14 |  |  |
| **Religion** |  |  |  |  |  |
| Christianity | 18 (22.0%) | 64 (78.0) | 82 | 8.223 | 0.016 |
| Islam | 18 (42.9%) | 24 (57.1%) | 42 |  |  |
| Traditional | 1(100%) | 0 (0) | 1 |  |  |
| **Sex** |  |  |  |  |  |
| Male | 21(36.2%) | 37(63.8%) | 58 | 2.267 | 0.095 |
| Female | 16(23.9%) | 51(76.1%) | 67 |  |  |
| **Class** |  |  |  |  |  |
| JSS | 21(33.9%) | 41(66.1%) | 62 | 1.077 | 0.299 |
| SSS | 16(25.4%) | 47(74.6%) | 63 |  |  |
| **Ethnicity** |  |  |  |  |  |
| Yoruba | 30(28.6%) | 75(71.4%) | 105 | 0.573 | 0.751 |
| Igbo | 6(33.3%) | 12(66.7%) | 18 |  |  |
| Hausa | 1(50.0%) | 1(50.0%) | 2 |  |  |
| **Live with** |  |  |  |  |  |
| Father | 1(7.7%) | 12(92.3%) | 13 | 4.677 | 0.197 |
| Mother | 28(26.1%) | 17(73.9%) | 23 |  |  |
| Both parent | 27(35.5%) | 49(64.5%) | 76 |  |  |
| Guardians | 3(23.1%) | 10(76.9%) | 13 |  |  |
| **Parents' Marital Status** |  |  |  |  |  |
| Never married | 1(25.0) | 3(75.0%) | 4 | 5.348 | 0.253 |
| Married | 28(31.5%) | 61(68.5%) | 89 |  |  |
| Divorced | 0 | 7(100%) | 7 |  |  |
| Separated | 7(41.2%) | 10(58.8%) | 17 |  |  |
| Widowed | 1(12.5%) | 7(87.5%) | 8 |  |  |
| **Work** |  |  |  |  |  |
| Yes | 17(30.4%) | 39(69.6%) | 56 | 0.028 | 0.867 |
| No | 20(29.0%) | 49(71.0%) | 69 |  |  |
| **Closeness to father** |  |  |  |  |  |
| Very close | 1 (20.0%) | 4(80.0%) | 5 | 2.959 | 0.938 |
| Close | 11(31.4%) | 24(68.6%) | 35 |  |  |
| Not close | 25(31.6%) | 54(68.4%) | 79 |  |  |
| Never had a relationship | 0 | 6(100%) | 6 |  |  |
| **Closeness to mother** |  |  |  |  |  |
| Very close | 14(56.0%) | 11(44.0%) | 25 | 12.620 | 0.006 |
| Close | 12(31.6%) | 26(68.4%) | 38 |  |  |
| Not close | 10(17.9%) | 46(82.1%) | 56 |  |  |
| Never had a relationship | 1(16.7%) | 5(83.3%) | 6 |  |  |

A higher percentage of those in junior secondary school (33.9%) compared to those in senior secondary school (25.4%) reported their experience of sexual violence, although this was not statistically significant. At the **relationship level**, closeness to the mother was significantly associated with reporting (χ² = 12.62, p = 0.006), whereas closeness to the father, living arrangements, and parental marital status were not significantly associated. Adolescents who reported being very close or close to their mothers were more likely to disclose experiences of sexual violence than those who were not close or had no relationship with their mothers.

## Discussion

Many adolescents face sexual violence in various contexts in which they interact with others without disclosing their experiences to anybody; this poses a challenge to their health and general well-being with immediate and long-term consequences. This study explored the prevalence and reporting of sexual violence among adolescents in Ibadan metropolis and factors associated with reporting. Around the time of data collection for this study, the COVID-19 pandemic restrictions made adolescents more vulnerable to sexual violence, with economic restrictions, school closures, and limited healthcare access increasing their susceptibility to abuses and limited support during lockdowns (18,37).

The mean age of participants was 14.6 ± 1.7 years, with most respondents in mid-adolescence comparable with reports from previous studies among in-school adolesscents (5,10). In this study, 46.7% of the respondents have had a romantic partner, and 26.4% currently have a romantic partner; this is in line with reports by previous report where 51.7% of the participants were in a relationship (38). Romantic relationships often start during the adolescent phase, increasing vulnerabilities and the risk of experiencing sexual violence (39).

This study employed the WHO definition of sexual violence and has a more comprehensive list, including non-contact forms. Almost half (42.5%) of the respondents had a lifetime experience of at least one act of sexual violence, while approximately one-third (34.7%) experienced sexual violence within the 12 months preceding this study. This is relatively high compared to the findings of 27.63% reported by Lasisi and Ozurumba 2021 among adolescents in Ilorin, Nigeria (10) and may be due to differences in the definition of sexual violence and the types of sexual violence assessed.

Unwanted sexual touching was the most commonly reported form of sexual violence in this study (18.3% lifetime prevalence and 25.6% twelve months prior), consistent with findings from other studies among adolescents in Nigeria, including Ajuwon. *et al.,* 2011 et al who reported a 22.7% prevalence (22). Rape and attempted rape were less frequently reported (lifetime 9.7% and 12 months prior 19.2 %) in this study, although their occurrence remains concerning, comparable with reports from other similar studies, including 18.8% forced sexual intercourse reported by Abreu et.al., 2025, among in-school adolescents in South West Nigeria (40).

Perpetrators of sexual violence were mostly individuals known to the adolescents, including romantic partners, friends, neighbours, and classmates. In the 12 months prior to the study, they included romantic partners (36.0%), friends (16.0%), neighbours (14.4%), classmates or schoolmates (12.8%). This is consistent with the findings of other studies (22,38). The most common locations where sexual violence occurred were victims’ homes (46.4%), perpetrators’ homes (20.0%), markets/shops (14.4%), schools (13.6%), and, to a lesser extent, other people’s homes (5.6%). It has also been documented in previous literature that sexual violence against adolescents is perpetrated by known people and in familiar settings (20,41). These findings underscore the need for proper monitoring of adolescents and for ensuring their safety regardless of the setting or location in which they are.

The consequences of sexual violence reported in this study extended beyond physical harm, with fear (43.2%), physical injury (24.0%), and guilt (20.8%) being the most frequently cited outcomes. These psychological consequences can contribute to delayed disclosure and may have long-term implications for adolescents’ mental health, well-being, and social functioning, as has been documented (27,42,43). Survivors of sexual violence thus require various services, such as educational, emotional, and psychosocial support, sexual and reproductive health services and treatment for physical injuries (3,44).

Many adolescent survivors of sexual violence do not report their experience of sexual violence to anyone for various reasons, and this is evident in the low rates of reporting recorded across different studies (26,45–47). In this study, only 29.6% of survivors reported their experience to anyone, similar to rates reported in previous studies: 22.2% (10), 18% (8), and 9% (5). This underreporting calls for immediate action as adolescents need to access timely help following an experience of sexual violence to mitigate the negative physical, mental, psychological, and social consequences that may occur as a result of their victimisation (5).

Some of the major factors, such as fear, shame, guilt, non-recognition of some acts of sexual violence, and threat to life, all contribute to non-disclosure by adolescents ((5,25). The most frequently reported barriers to reporting in this study were feeling it was their fault (46.6%), not thinking it was a problem (31.8%), being afraid of getting in trouble (30.7%), and being embarrassed for themselves or their family (27.3%). Among those who chose to report, many do not seek formal help; this also has implications for both research and programs, as most of the informal disclosures may not provide adequate support to the survivors of sexual violence.

There was a significant association between age and closeness to the mother and reporting of sexual violence. Younger adolescents and those who reported closer relationships with their mothers were more likely to disclose their experience. However, there was no significant association between reporting and sex, closeness to father, engagement in trade for financial remuneration, or class.

### 5.2 Conclusion

This study shows that sexual violence is common among in-school adolescents in Ibadan, yet reporting remains low. Survivors rarely speak up or seek help, which has consequences for their health and well-being. The most commonly experienced act of sexual violence was unwanted sexual touching, and the perpetrators were often known, and incidents mostly occurred in familiar settings. Many of the adolescents did not report their experience due to fear, shame, guilt, and threats. These findings highlight the need for holistic, targeted interventions to address sexual violence among adolescents and strengthen response systems.

### 5.3 Recommendations

This study highlights the need to engage various stakeholders, including adolescents, parents, local communities, public health professionals, governments, and non-governmental organisations, to strengthen prevention and response to sexual violence. Parents, caregivers, and the broader community should be trained on responding to sexual violence and how to handle disclosure by adolescents, to avoid blaming and shaming the survivors. Adolescents should be assured of their safety, and schools should integrate comprehensive information on sexual violence, consent, and available reporting pathways into curricula.

At the systems level, there should be increased awareness of available laws, policies and response structures within communities. The police force and other law enforcement agencies should be trained to reassure victims and offer protection when necessary, and cases of successful prosecution of sexual offenders should be publicised and cited as examples to reassure survivors that they can get justice, to encourage reporting.

### 5.4 Limitations of the study

The study was limited to only in-school adolescents present in the schools during recruitment. Therefore, the study may not apply to adolescents who are out of school, and their experiences may differ. Some participants needed assurance before revealing sensitive information; this was addressed by assuring them of anonymity, building trust, and ensuring that data were collected in a secure location within the school. Due to the sensitive nature of this topic, underreporting of prevalence cannot be ruled out entirely, despite the provision of privacy during data collection. Given the cross-sectional nature of the data, the reported association may be influenced by other factors.

## Abbreviations

SV: Sexual Violence

## Declarations

### Ethics approval

This study involves human participants, and Ethical approval was obtained from the Oyo State Ethics Review Committee (Reference Number: AD 13/479/4485^A^). Participants provided informed consent before participating in the study.

### Consent for Publication

Not Applicable

### Availability of data and materials

The datasets used and/or analysed during the current study are available from the corresponding author on reasonable request.

### Author Contributions

HO conceived the research idea, designed the study, collected data, analysed the data, and drafted the manuscript. AO supervised the research and contributed to the writing of the manuscript. Both authors agreed on the final copy of the manuscript. HO is the guarantor for the publication.

## Acknowledgements

Not applicable

